# Medical Doctors’ Awareness, Perception, and Attitude towards COVID-19 in Bangladesh: A Cross sectional study

**DOI:** 10.1101/2020.05.14.20101659

**Authors:** Sadia Biswas Mumu, Most. Nasrin Aktar, Zabun Nahar, Shahana Sharmin, Md Shaki Mostaid

**Affiliations:** Department of Pharmacy, North South University, Dhaka-1229, Bangladesh; Department of Pharmacy, University of Asia Pacific, Dhaka-1215, Bangladesh; Department of Pharmacy, Brac University, Dhaka-1212, Bangladesh

**Keywords:** COVID-19, Medical doctors, awareness, perception, attitude, Bangladesh.

## Abstract

**Objective:** COVID-19 has emerged as a pandemic and during the first week of May Bangladesh has reported more than 10,000 cases. A lack of awareness and poor understanding of the disease may result in rapid transmission of the disease in Bangladesh. This study aimed to investigate the awareness, perception, and attitude towards COVID-19 among Bangladeshi medical doctors.

**Method:** This cross sectional, web-based study was conducted with the help of an online questionnaire and sent to the doctors which comprised of a series of questions regarding demographics of the participants, symptoms and incubation period of COVID-19, mode of transmission, measures to prevent transmission, availability of training and personal protective equipment in Bangladeshi hospitals, and attitude of doctors towards the treatment of suspected patients with COVID-19.

**Results:** Of 800 medical doctors, a total 545 completed the survey (response 68.13%). Among the participants, 52.3% were females, 72.8% were below 30 years of age, and majority (52.8%) were working outside the cities in the villages and rural areas. A total of 404 (74.1%) doctors reported the correct incubation period of COVID-19. Majority doctors were aware of the symptoms with mode of transmission of COVID-19, measures to prevent hospital transmission, along with ways of identifying suspected patients with COVID-19. However, more than 90% of the doctors reported of inadequate intensive care unit and ventilator facilities along with extreme scarcity of personal protective equipment in the hospitals. 65.7% doctors prefer avoid working with a COVID-19 patient and more than 50% doctors have expressed that they would send the suspected COVID-19 patients to designated hospitals without providing treatment.

**Conclusion:** The health authorities should take appropriate training measures to increase the awareness of the medical doctors along with providing sufficient amount of personal protective equipment for the medical doctors and supporting staff before deploying them in hospitals.

## Introduction

The COVID-19 disease has become a pandemic affecting almost all the countries in the world. Since December, 2019 and as of 24^th^ April, 2020 approximately 2,719,897 COVID-19 patients have been reported including 187,705 deaths [1]. COVID-19 seems more contagious than its counterparts SARS (severe acute respiratory syndrome) and MARS (middle-east acute respiratory syndrome) and this highly infective disease may transmit from person to person contact via respiratory droplets [2], fecal-oral transmission [3], vertical transmission [4]. Majority of patients with this disease have experienced high fever with dry cough while others showed symptoms like shortness of breath, fatigue, muscle pain along with confusion, headache, sore throat, diarrhea, and vomiting [5]. On the contrary, up to 25% of COVID-19 patients remain asymptomatic and do not manifest any clear clinical symptoms are therefore remains unidentified but carry a high risk of transmitting the infection to their family members and society [6, 7]. So, it is difficult to identify and quarantine these patients on time. Moreover, patients in the recovering phase of COVID-19 are also potential sources of transmission. As a result, social and physical distancing has been suggested as an efficient tool to control the transmission of COVID-19 [8]. Frequent washing of hands using alcohol-based hand sanitizer or soap and water, use of face mask and covering mouth and nose with elbow or disposable tissues while coughing and sneezing are recommended measures to prevent the spread of COVID-19 [9].

### COVID-19 and Bangladeshi healthcare perspective

Recently, this novel coronavirus has undergone community transmission in Bangladesh. As of 26 April 2020, there are 5416 confirmed COVID-19 cases in Bangladesh, including 122 patients recovered and 145 related deaths reported by the Institute of Epidemiology, Disease Control & Research, [10]. Although people of all ages are generally susceptible to this infective disease but those who are in close contact with patients with COVID-19 including health care workers and patients in the hospital are at excessive risk of getting infected with this deadly virus.

Bangladesh government claims that it started taking preparation to control the COVID-19 pandemic in the country since January 2020 based on the guidelines of the World Health Organization (WHO). While the first COVID-19 patient was detected on March 08, 2020, the national guidelines for the prevention of COVID-19 in health care setting was first published by the Directorate General of Health Services under the Ministry of Health and Family Welfare on March 19, 2020 [11] and the national guidelines on clinical management of COVID-19 was published on March 30, 2020 [12].

It is of utmost importance to have proper knowledge, awareness and perceptions regarding COVID-19 for all health care workers. Proper training of all the health care workers especially medical doctors is mandatory before posting them to treat COVID-19 patients in designated hospitals as well is all other clinics during the pandemic situation. According to the ministry of health and family welfare HRH data sheet 2014, there are 75,514 registered medical doctors and 38,452 registered diploma nurses in Bangladesh [13] and as of 20^th^ April, 2020 only 3,625 medical doctors and 1,314 nurses have received video training on COVID-19 hospital management and hospital infection prevention and control [14]. So, there is a clear shortage of training on COVID-19 for health care workers in Bangladesh which may result in lack of knowledge with incorrect perception and attitude regarding COVID-19 among health care workers especially medical doctors.

Protection of health care workers is vital as they are at the highest risk of getting infected with COVID-19 due to continuous exposure to the infected patients at various clinics and hospitals [15]. Adequate amount of medical resources is required during this pandemic with adequate healthcare infrastructures. Proper personal protective equipment (PPE) is also compulsory to prevent the transmission of the virus from patients to doctors and allied health care workers. Inadequate personal protection, long time exposure to the infected patients, lack of proper PPE, inadequate training are some reasons for the rapid spread of the COVID-19 infection among health care workers [16].

With all the recommendation and management protocols provided by the World Health Organization and government initiatives, all the medical doctors are expected to gain proper knowledge, awareness, and clear perceptions about COVID-19. Hence, our study aimed to assess the level of awareness, perception and attitude regarding COVID-19 and infection control among the Bangladeshi doctors who are working at various hospitals at both city and rural areas.

## Materials and Methods

### Study Population

The study population comprised of registered medical doctors who are currently working in Bangladesh, both in private hospitals and public hospitals including armed forces. This survey was conducted in April 2020. Here we developed an online questionnaire using Google Forms that was used to collect the data.

The sample of doctors was selected through personal contact from different hospitals as well as from Facebook groups created by the registered medical doctors from Bangladesh Medical Association (BMA). Some of them were Government employees while majority were working in private hospitals and clinics in different parts of Bangladesh. The Google Form was made accessible through a link and was advertised in different Facebook groups of doctors. Although there were numerous groups, four groups were randomly chosen: Bangladesh Doctors’ Foundation (BDF), Doctors Society of Bangladesh (DSF), Doctors’ Club, Doctors’ Association of Bangladesh (DAB).

Within the groups, 800 doctors were randomly selected and invited to participate in the study by their Facebook profiles. However, each participant randomly selected was contacted individually to make sure that he/she was a doctor and currently working in Bangladesh.

Confidentiality of information was maintained throughout the study by making the participants’ information and responses anonymous. All the doctors participated voluntarily and had to sign an electronic consent form. The study was conducted following the Helsinki Declaration as revised in 2013 [17] and the Checklist for Reporting Results of Internet E-Surveys (CHERRIES) guidelines [18]. Ethical approval was obtained from the Institutional Review Board at BRAC University.

### Study design and Data Collection

The survey questions were developed based on reviewing relevant literature and the international guidelines [6, 19–21]. The questionnaire was designed in English and it contained structured multiple-choice questions divided into sections related to socio demographic and profession-related characteristics, the knowledge of doctors about COVID-19 (symptoms, incubation period, and mode of transmission), doctors’ awareness of measures for infection prevention of COVID-19 in hospitals, doctors’ viewpoints on hospital facilities and availability of personal protective equipment in Bangladesh, and their attitudes toward treatment of patients with COVID-19.

### Data Analysis

Data were analyzed using SPSS. Descriptive statistical analysis was used to describe items included in the survey. Mean and standard deviation or median and range were used to describe the continuous variables, and percentages were used to describe the categorical data.

## Results

### Demographics of the participant medical doctors

A total of 545 (males 249 and females 296) doctors participated in this study. Majority doctors (397) were below 30 years of age, while only 15 doctors were more than 40 years old. The age range was 26–65 with a median of 42.3 years. Among the participants, 274 (50.3%) doctors were from the private sectors, 171 doctors (31.4%) working in public hospitals. Moreover, 275 (50.3%) doctors were practicing in district towns, 185 (33.9%) from the capital city Dhaka, and 42 (7.7%) from rural areas and villages. The details of the demographics are given in Table 1.

**Table 1.** Demographics of the medical doctors enrolled in the study (n = 545). **Variable Doctors, n (%)**

| **Variable** | **Doctors, n (%)** |
| --- | --- |
| **Gender** |  |
| Male | 249 (45.7) |
| Female | 296 (54.3) |
| **Age** |  |
| <30 | 397 (72.8) |
| 31-40 | 133 (24.4) |
| ≥ 40 | 15 (2.8) |
| **Educational level** |  |
| Only MBBS | 425 (78) |
| MBBS with Post Graduation Diploma | 65 (11.9) |
| MD (Res)/PhD/Higher Degrees | 55 (10.1) |
| **Years of practice** |  |
| <5 | 416 (76.3) |
| 5-10 | 94 (17.3) |
| >10 | 35 (6.4) |
| **Working Sector** |  |
| Public sector only | 171 (31.4) |
| Private sector only | 274 (50.3) |
| Armed force | 13 (2.4) |
| Both private and public | 87 (15.9) |
| **Work station** |  |
| Dhaka Metropolitan area | 185 (33.9) |
| Outside Dhaka (district level) | 275 (50.5) |
| Rural areas (Village) | 42 (7.7) |
| Practices both inside and outside Dhaka Metropolitan area | 43 (7.9) |
| **Working shift** |  |
| Day shift | 515 (94.5) |
| Night shift | 30 (5.5) |

### Awareness of COVID-19 infection among the medical doctors in Bangladesh

Majority of the medical doctors in Bangladesh are aware on many aspects of the COVID-19 infection. 404 (74.1%) doctors reported the correct incubation period (1–14 days) of COVID-19. But it is a concern that almost 25% are still not aware of this critical information. When asked about the symptoms, over 90% of the doctors pointed out fever, dry cough, shortness of breath, and sore throat (Table 2). Runny nose, and diarrhea were reported by more than 50% of the doctors. Meanwhile, 394 (72.3%) doctors said that COVID-19 could be asymptomatic. Only a small percentage expressed that skin rash, vomiting, and joint or muscle pain are symptoms of this disease.

**Table 2.** Doctors’ awareness about incubation period, symptoms, and mode of transmission of the coronavirus disease infection (n = 545).

| **Variable** | **Doctors, n (%)** |
| --- | --- |
| **Incubation period (days)** |  |
| 1-14 | 404 (74.1) |
| 2-7 | 8 (1.5) |
| 7-14 | 82 (15.1) |
| 7-21 | 51 (9.4) |
| **Symptoms of the COVID-19 infection** |  |
| Fever | 540 (99.1) |
| Dry cough | 530 (97.3) |
| Shortness of breath | 530 (97.3) |
| Diarrhea | 434 (79.7) |
| Vomiting | 147 (27.0) |
| Runny nose | 283 (52.0) |
| Sore throat | 494 (90.7) |
| Red eyes | 126 (23.1) |
| Skin rash | 71 (13.0) |
| Joint or muscle pain | 207 (37.9) |
| May present with no symptoms | 394 (72.3) |
| **Mode of transmission** |  |
| Coughing and sneezing | 529 (97.1) |
| Embrace and hand shaking | 470 (86.2) |
| Touching surfaces such as doorknobs and tables | 464 (85.1) |
| Food | 96 (17.6) |
| Body fluids (blood, saliva, sexual fluids) | 161 (29.6) |

The medical doctors could correctly point out the mode of transmission of COVID-19 (Table 2). When they were asked about how to identify suspected COVID-19 patients, 527 (96.7%) mentioned presence of fever, coughing, sneezing along with difficulty in breathing, 530 (97.3%) expressed close contact with a COVID-19 patient, 504 (92.5%) mentioned coming from an area already infected with COVID-19.

### Doctors awareness of measures for preventing the transmission of COVID-19 in hospitals

Bangladeshi doctors were found to be well aware of the necessary measures that are required for preventing the transmission of COVID-19 in hospital settings. Most of the doctors (above 90%) emphasized on frequent cleaning of hands with soap and water or the use of alcohol based hand sanitizers, use of proper personal protective equipment, routine cleaning of surfaces with confirmed or suspected patients. Moreover, they also pointed out putting face masks on suspected COVID-19 patients and avoid of transporting patients unless in emergency. The percentage of doctors who supported the above-mentioned measures along with other specific measures are shown in Table 3.

**Table 3.**
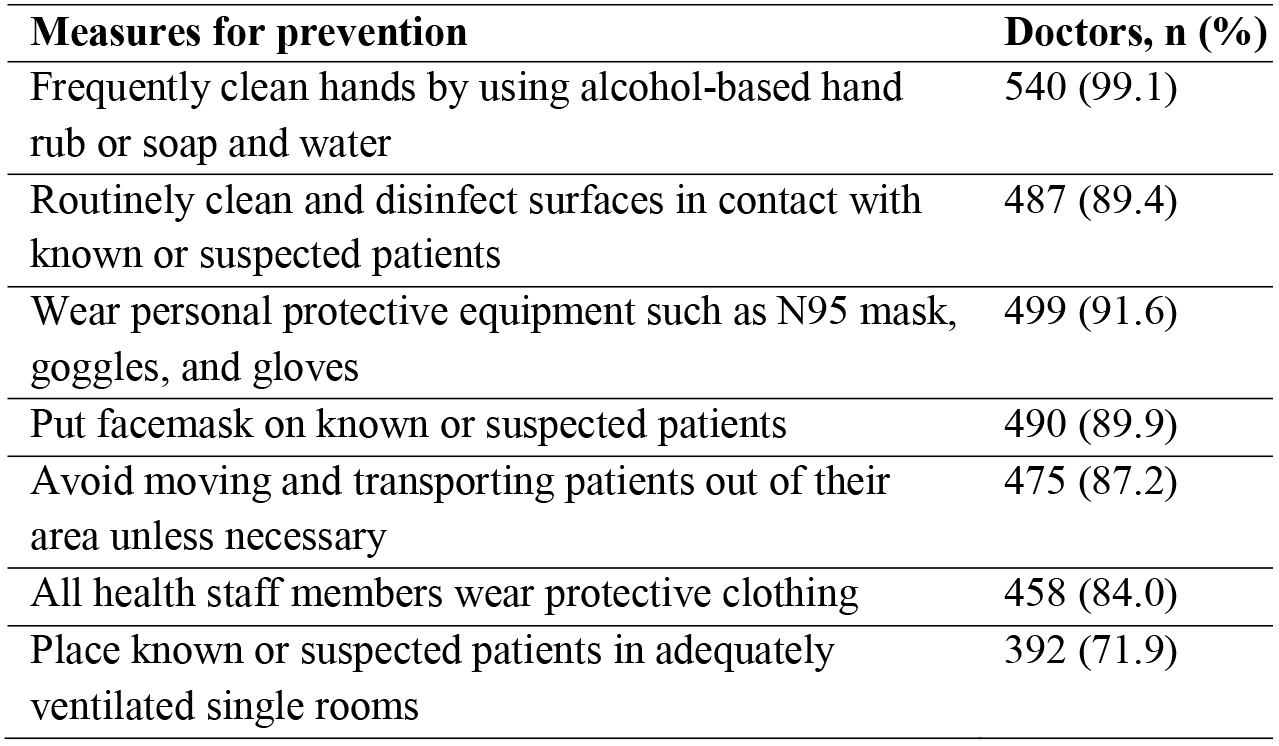
Doctors’ awareness of measures for the prevention of coronavirus disease transmission in hospitals (n = 545).

| **Measures for prevention** | **Doctors, n (%)** |
| --- | --- |
| Frequently clean hands by using alcohol-based hand rub or soap and water | 540 (99.1) |
| Routinely clean and disinfect surfaces in contact with known or suspected patients | 487 (89.4) |
| Wear personal protective equipment such as N95 mask, goggles, and gloves | 499 (91.6) |
| Put facemask on known or suspected patients | 490 (89.9) |
| Avoid moving and transporting patients out of their area unless necessary | 475 (87.2) |
| All health staff members wear protective clothing | 458 (84.0) |
| Place known or suspected patients in adequately ventilated single rooms | 392 (71.9) |

### Perception of COVID-19 among Bangladeshi medical doctors

A vast majority of doctors (515, 94.5%) perceived that COVID-19 is very dangerous and 498 (91.9%) doctors believed that it is a serious public health issue. Majority of the doctors 508 (93.2%) agree that at present, lockdown of cities is the best possible method for preventing the transmission of COVID-19.

### Doctors’ viewpoint on hospital facilities and availability of PPE regarding COVID-19 patient care in Bangladesh

According to our survey, most of the doctors’ reported that the patient care facilities and availability of PPEs for the proper care of COVID-19 patients in Bangladesh is very low and nowhere near sufficient compared to the overwhelming requirement at this current pandemic situation. More than 90% of the doctors’ reported that there were not enough ventilators, and intensive care units (ICUs) in the hospitals. Moreover, 499 (91.5%) doctors reported that they were not provided with sufficient amount of high quality PPEs. In addition to that doctors were not provided with training on COVID-19 and rational use of PPEs before being posted in hospitals. The details percentage are given inTable 4.

**Table 4.**
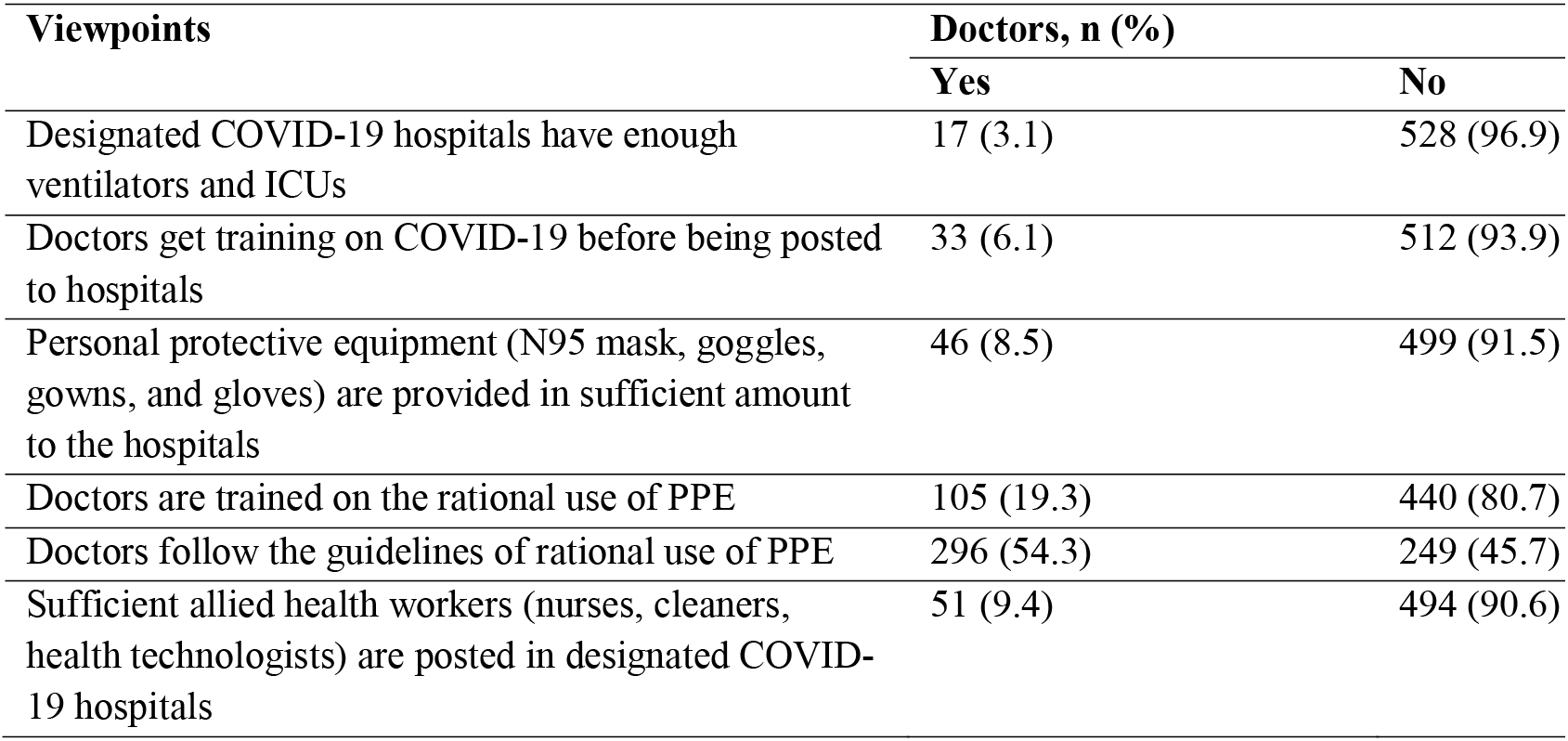
Doctors’ viewpoint on Hospital facilities and availability of PPE regarding COVID-19 patient care in Bangladesh.

| Viewpoints | Doctors, n (%) |  |
| --- | --- | --- |
|  | Yes | No |
| Designated COVID-19 hospitals have enough ventilators and ICUs | 17 (3.1) | 528 (96.9) |
| Doctors get training on COVID-19 before being posted to hospitals | 33 (6.1) | 512 (93.9) |
| Personal protective equipment (N95 mask, goggles, gowns, and gloves) are provided in sufficient amount to the hospitals | 46 (8.5) | 499 (91.5) |
| Doctors are trained on the rational use of PPE | 105 (19.3) | 440 (80.7) |
| Doctors follow the guidelines of rational use of PPE | 296 (54.3) | 249 (45.7) |
| Sufficient allied health workers (nurses, cleaners, health technologists) are posted in designated COVID-19 hospitals | 51 (9.4) | 494 (90.6) |

**Table 5.** Doctors’ attitude regarding treatment of patients with COVID-19 **Measures of attitude**.

| Measures of attitude | Doctors, n (%) |
| --- | --- |
| The symptoms of COVID-19 often resolve with time | 409 (75.1) |
| Patients should wear masks and gloves while coming to the hospital | 461 (84.6) |
| Prefer avoid working with a COVID-19 patient | 358 (65.7) |
| If a patient comes with coughing and sneezing comes for treatment: |  |
| a. Treat the patient and advise them to go to the COVID-19 designated hospitals | 241 (44.2) |
| b. Refer the patient to COVID-19 designated hospitals without treating them | 273 (50.1) |
| c. Refuse treating the patient and ask them to leave the clinic | 31 (5.7) |

### Bangladeshi doctors’ attitude towards the treatment of COVID-19 patients

More than 75% of the doctors (409) reported that the symptoms of COVID-19 resolve gradually and do not require any medication. 461 (84.6%) doctors have expressed that patients should wear masks and gloves while coming to the hospital at this time of pandemic to reduce the risk of transmission. However, 358 (65.7%) doctors prefer to avoid working with a COVID-19 patient.

When asked about their approach when a patient comes with coughing and sneezing, half of the doctors (273, 50.1%) mentioned that they would refer the patients to the designated COVID-19 hospitals without any treatment. 241 (44.2%) reported that they would give treatment but advise them to go to the COVID-19 designated hospitals and 31 (5.7%) mentioned that they would refuse treatment.

The doctors were also aware of the medications that are being used on experimental basis to treat COVID-19 in different countries and 369 (67.7%) mentioned that they would use the combination of Hydroxychloroquine and Azithromycin. 288 (52.9%) prefer to use Favipiravir and 132 (24.2%) prefer Remdesivir. 141 (25.9%) reported that there would not use any of these medicines on an experimental basis.

## Discussion

Our study provides an overview on the level of awareness, perception and attitude of Bangladeshi medical doctors regarding COVID-19 at the time of pandemic in 2020. Moreover, it gives an insight on the hospital facilities and availability of PPE for doctors in Bangladesh who are at the forefront to fight this contagious disease.

Majority of the doctors participated in this study were below 30 years of age. Moreover, the number of the female doctors participated were higher than the male doctors (54.3% vs 45.7%) which is in support of the fact that the number of female doctors (52%) in Bangladesh is slightly higher than the number of male doctors (48%) [22]. Most of the doctors (74.1%) in our study had the correct knowledge about the incubation period (up to 14 days) of COVID-19 [23], but it becomes a concern that almost 25% are still not aware of this critical information. It is essential to know the right incubation period of COVID-19 because of its role in determining the safe period to treat suspected patients [23].

Prime symptoms of COVID-19 include fever, dry cough, shortness of breath, sore throat, diarrhea [24, 25]. Most of the doctors in our study could identify the major symptoms of COVID-19. Knowledge of the symptoms would help the doctors to identify the suspected patients and take the necessary steps to prevent spreading of the disease to other patients, and health care workers in the hospital.

Doctors’ awareness of measures for preventing the transmission of COVID-19 was up to the satisfactory level according to our study. The most recommended measure is frequent sanitization of hands by using alcohol-based hand rub or soap and water along with wearing personal protective equipment while at the hospital [9]. At present, there is no specific treatment for COVID-19. Measures like hand sanitization and social distancing to control infection transmission, testing of all suspected patients, isolation, mandatory quarantine are some of the steps taken by several countries including Bangladesh. Providing adequate ICU and ventilation support to the critical patients with COVID-19 is also an important step to reduce the number of deaths [24].

Regarding the perception of COVID-19, 94.5% of our participants believe that COVID-19 is very dangerous and lockdown, quarantine, social distancing, isolation of the patients are effective ways to prevent the transmission of infection. Quarantine had been used to contain or minimize infectious disease outbreaks, particularly in the developing world [26, 27]. However, it can also be very effective in protecting or restoring public health. As a result, with the outbreak of the COVID-19, various quarantine policies have been implemented by most of countries in order to reduce transmission of the disease.

Doctors need sufficient amount of proper personal protective equipment to protect themselves from being infected while providing treatment to the COVID-19 patients. Doctors have also reported that the amount of ICU and ventilator facilities in Bangladeshi hospitals are extremely low to meet the need of its large population. This is supported by the data provided by Directorate General of Health services of Bangladesh Government which reports that only 3302 oxygen cylinders and 49 ventilators are available for COVID-19 treatment till 08 May 2020. Moreover, only 757 physicians and 693 nurses have been posted for COVID-19 treatment all over Bangladesh [28], which is insufficient by all means. A somewhat similar scenario was observed even in developed countries where the hospitals fell short of providing ventilator and ICU support to meet the overwhelming demand of the COVID-19 patients [29]. Moreover, doctors have expressed that they were not provided with sufficient training and PPEs before being deployed in the hospitals in Bangladesh. As, a result more than 250 doctors got infected with COVID-19 [30] and thus got isolated which is creating extra burden on an already fragile health care system.

Approximately, 75% doctors believe that the symptoms of COVID-19 resolve with time and so the patients should remain at home and get symptomatic treatments. 84.6% doctors have expressed that all patients should use mask and gloves while coming to the hospital for any sort of emergency treatment to prevent nosocomial transmission of COVID-19. However, 65.7% doctors (n=358) would prefer not working with a COVID-19 patient which may be due to the lack of PPEs and insufficient training which increases their risk of being infected. If a patient comes with coughing and sneezing, almost half of the doctors (n=273, 50.1%) would suggest them to go to the designated COVID-19 hospitals, 44.2% (n= 241) would provide treatment with an advice to go to the COVID-19 designated hospitals and 5.7% (n= 31) would totally refuse to provide treatment. This scenario shows an overall tensed condition that exists among the medical doctors regarding COVID-19 in Bangladesh.

Our study had some limitations. The number of participants were of moderate size. This might be due to a relatively short period of data collection. Many of the doctors might be occupied with getting latest COVID-19 information on television and internet while taking care of their nearest and dearest ones. Also, they might be busy to their duties in this current pandemic situation of COVID-19. So, only the medical doctors who were available on the social media within the short period of data collection, participated in this survey. This could result in sampling error and selection bias and so caution should be taken before generalization of our results.

## Conclusion

To conclude, Bangladeshi doctors were well aware of COVID-19 symptoms, mode of transmission, infection control and measures for preventing the transmission of COVID-19 in hospitals which may be attributed to their own efforts of acquiring knowledge by themselves through television, internet and other sources rather than government initiatives. But considering the severity of COVID-19 pandemic situation they were not provided with sufficient training and quality PPEs which are very essential for protecting the doctors while treating the COVID-19 patients. So, the government of Bangladesh should try to increase hospital facilities, provide training, and supply enough PPEs to all medical doctors and supporting staff for proper management of COVID-19.

## Data Availability

All data has been provided in the manuscript and related files

## Acknowledgements

The authors are thankful to the medical doctors who participated in the survey voluntarily. The authors would also like to thank the collaborators of the Bangladesh Medical Association who helped to carry out the research work.

## References

1. Coronavirus disease 2019 (COVID-19) Situation Report – 96 [Internet]. World Health Organization. 2020 [cited 26 April 2020]. Available from: https://www.who.int/emergencies/diseases/novel-coronavirus-2019/situation-reports/.

2. Rothan HA, Byrareddy SN. The epidemiology and pathogenesis of coronavirus disease (COVID-19) outbreak. J Autoimmun. 2020;109:102433. doi: 10.1016/j.jaut.2020.102433. PubMed PMID: 32113704; PubMed Central PMCID: PMCPMC7127067.

3. Gu J, Han B, Wang J. COVID-19: Gastrointestinal Manifestations and Potential Fecal-Oral Transmission. Gastroenterology. 2020. doi: 10.1053/j.gastro.2020.02.054. PubMed PMID: 32142785; PubMed Central PMCID: PMCPMC7130192.

4. Chen H, Guo J, Wang C, Luo F, Yu X, Zhang W, et al. Clinical characteristics and intrauterine vertical transmission potential of COVID-19 infection in nine pregnant women: a retrospective review of medical records. Lancet. 2020;395(10226):809–15. doi: 10.1016/S0140-6736(20)30360-3. PubMed PMID: 32151335; PubMed Central PMCID: PMCPMC7159281.

5. Wu YC, Chen CS, Chan YJ. The outbreak of COVID-19: An overview. J Chin Med Assoc. 2020;83(3):217–20. doi: 10.1097/JCMA.0000000000000270. PubMed PMID: 32134861; PubMed Central PMCID: PMCPMC7153464.

6. Chan JF, Yuan S, Kok KH, To KK, Chu H, Yang J, et al. A familial cluster of pneumonia associated with the 2019 novel coronavirus indicating person-to-person transmission: a study of a family cluster. Lancet. 2020;395(10223):514–23. doi: 10.1016/S0140-6736(20)30154-9. PubMed PMID: 31986261; PubMed Central PMCID: PMCPMC7159286.

7. Bai Y, Yao L, Wei T, Tian F, Jin DY, Chen L, et al. Presumed Asymptomatic Carrier Transmission of COVID-19. JAMA. 2020. doi: 10.1001/jama.2020.2565. PubMed PMID: 32083643; PubMed Central PMCID: PMCPMC7042844.

8. Prem K, Liu Y, Russell TW, Kucharski AJ, Eggo RM, Davies N, et al. The effect of control strategies to reduce social mixing on outcomes of the COVID-19 epidemic in Wuhan, China: a modelling study. Lancet Public Health. 2020. doi: 10.1016/S2468-2667(20)30073-6. PubMed PMID: 32220655; PubMed Central PMCID: PMCPMC7158905.

9. Desai AN, Patel P. Stopping the Spread of COVID-19. JAMA. 2020. doi: 10.1001/jama.2020.4269. PubMed PMID: 32196079.

10. IEDCR. Dhaka, Bangladesh: Institute of Epidemiology, Disease Control and Research; 2020 [cited 2020 27 April]. Available from: http://www.iedcr.gov.bd/.

11. DGHS. National Guideline for Health Care Provider On Infection Prevention and Control of COVID-19 pandemic in Healthcare Setting: Directorate General of Health Services, Ministry of Health and Family Welfare, Government of the Peoples’ Republic of Bangladesh; 2020 [cited 2020 27 April]. Available from: https://dghs.gov.bd/images/docs/Guideline/IPC_Module_for_COVID-19_for_frontline_HCW_20.3.2020.pdf.

12. DGHS. National Guidelines on Clinical Management of Coronavirus Disease 2019 (Covid-19) Dhaka: Government of the Peoples’ Republic of Bangladesh; 2020 [cited 2020 27 April]. Available from: https://corona.gov.bd/documents/COVID_Guideline_V4.0..30.3.2020.pdf.

13. MoHFW. HRH Data Sheet 2014 of MOHFW: Ministry of Health and Family Welfare, Government of the Peoples’ Republic of Bangladesh; 2015 [cited 2020 28 April 2020]. Available from: http://www.mohfw.gov.bd/index.php?option=com_content&view=article&id=112.

14. Coronavirus disease (COVID-2019) Bangladesh situation reports-08 [Internet]. World Health Organization. 2020 [cited 28 April 2020]. Available from: https://www.who.int/docs/default-source/searo/bangladesh/covid-19-who-bangladesh-situation-reports/who-ban-covid-19-sitrep-08.pdf?sfvrsn=a108826d_4.

15. Lancet T. COVID-19: protecting health-care workers. Lancet. 2020;395(10228):922. doi: 10.1016/S0140-6736(20)30644-9. PubMed PMID: 32199474; PubMed Central PMCID: PMCPMC7138074.

16. Wang J, Zhou M, Liu F. Reasons for healthcare workers becoming infected with novel coronavirus disease 2019 (COVID-19) in China. J Hosp Infect. 2020. doi: 10.1016/j.jhin.2020.03.002. PubMed PMID: 32147406; PubMed Central PMCID: PMCPMC7134479.

17. World Medical A. World Medical Association Declaration of Helsinki: ethical principles for medical research involving human subjects. JAMA. 2013;310(20):2191–4. doi: 10.1001/jama.2013.281053. PubMed PMID: 24141714.

18. Eysenbach G. Improving the quality of Web surveys: the Checklist for Reporting Results of Internet ESurveys (CHERRIES). J Med Internet Res. 2004;6(3):e34. doi: 10.2196/jmir.6.3.e34. PubMed PMID: 15471760; PubMed Central PMCID: PMCPMC1550605.

19. WHO. Clinical management of severe acute respiratory infection when COVID-19 is suspected: World Health Organization; 2020 [cited 2020 3 May]. Available from: https://www.who.int/publications-detail/clinical-management-of-severe-acute-respiratory-infection-when-novel-coronavirus-(ncov)-infection-is-suspected.

20. Liu J, Liao X, Qian S, Yuan J, Wang F, Liu Y, et al. Community Transmission of Severe Acute Respiratory Syndrome Coronavirus 2, Shenzhen, China, 2020. Emerg Infect Dis. 2020;26(6). doi: 10.3201/eid2606.200239. PubMed PMID: 32125269.

21. CDC. Symptoms of Coronavirus: Centers for Disease Control and Prevention; 2020 [cited 2020 29 March]. Available from: https://www.cdc.gov/coronavirus/2019-ncov/symptoms-testing/symptoms.html.

22. Hossain P, Das Gupta R, YarZar P, Salieu Jalloh M, Tasnim N, Afrin A, et al. ‘Feminization’ of physician workforce in Bangladesh, underlying factors and implications for health system: Insights from a mixed-methods study. PLoS One. 2019;14(1):e0210820. doi: 10.1371/journal.pone.0210820. PubMed PMID: 30633775; PubMed Central PMCID: PMCPMC6329528

23. Lauer SA, Grantz KH, Bi Q, Jones FK, Zheng Q, Meredith HR, et al. The Incubation Period of Coronavirus Disease 2019 (COVID-19) From Publicly Reported Confirmed Cases: Estimation and Application. Ann Intern Med. 2020. doi: 10.7326/M20-0504. PubMed PMID: 32150748; PubMed Central PMCID: PMCPMC7081172.

24. Wang D, Hu B, Hu C, Zhu F, Liu X, Zhang J, et al. Clinical Characteristics of 138 Hospitalized Patients With 2019 Novel Coronavirus-Infected Pneumonia in Wuhan, China. JAMA. 2020. doi: 10.1001/jama.2020.1585. PubMed PMID: 32031570; PubMed Central PMCID: PMCPMC7042881.

25. Yang X, Yu Y, Xu J, Shu H, Xia J, Liu H, et al. Clinical course and outcomes of critically ill patients with SARS-CoV-2 pneumonia in Wuhan, China: a single-centered, retrospective, observational study. Lancet Respir Med. 2020. doi: 10.1016/S2213-2600(20)30079-5. PubMed PMID: 32105632; PubMed Central PMCID: PMCPMC7102538.

26. Kucharski AJ, Camacho A, Flasche S, Glover RE, Edmunds WJ, Funk S. Measuring the impact of Ebola control measures in Sierra Leone. Proc Natl Acad Sci U S A. 2015;112(46):14366–71. doi: 10.1073/pnas.1508814112. PubMed PMID: 26460023; PubMed Central PMCID: PMCPMC4655521.

27. Giubilini A, Douglas T, Maslen H, Savulescu J. Quarantine, isolation and the duty of easy rescue in public health. Dev World Bioeth. 2018;18(2):182–9. doi: 10.1111/dewb.12165. PubMed PMID: 28922559; PubMed Central PMCID: PMCPMC6001516.

28. DGHS. Government of the Peoples’ Republic of Bangladesh; 07 May 2020 [cited 2020 08 May]. Available from: http://dashboard.corona.gov.bd/.

29. Tanne JH, Hayasaki E, Zastrow M, Pulla P, Smith P, Rada AG. Covid-19: how doctors and healthcare systems are tackling coronavirus worldwide. BMJ. 2020;368:m1090. doi: 10.1136/bmj.m1090. PubMed PMID: 32188598.

30. Mahmud F. Hundreds of doctors in Bangladesh infected with coronavirus Qatar: Al Jazeera; 24 April 2020 [cited 2020 08 May]. Available from: https://www.aljazeera.com/news/2020/04/hundreds-doctors-bangladesh-infected-coronavirus-200423080515266.html.

